# Performance of a large language model (ChatGPT-3.5) for Pooled Cohort Equation estimation of atherosclerotic cardiovascular disease risk

**DOI:** 10.1101/2023.08.11.23293957

**Authors:** Ben J. Marafino, Vincent X. Liu

**Affiliations:** Kaiser Permanente Division of Research, 2000 Broadway, Oakland, CA 94612

## Abstract

Despite demonstrated facility for arithmetic and other quantitative tasks, the performance of ChatGPT and other large language models for clinical risk calculation have yet to be assessed. Using synthetic patient data, this preliminary study aimed to assess the calibration, reproducibility, and potential for sociodemographic bias of ChatGPT-derived Pooled Cohort Equation (PCE) scores of atherosclerotic cardiovascular disease risk as compared to true scores. We found that ChatGPT-derived PCE scores, despite being moderately associated with the true PCE scores, displayed poor calibration with respect to true PCE scores, and exhibited instability between repeated rounds of prompting, suggesting lack of reproducibility. Moreover, ChatGPT-derived PCE scores also appeared inappropriately sensitive to contextual indicators of the sociodemographic status of the synthetic patients in this study. Further work is needed to confirm these results, and to assess performance on a wider variety of prompts as well as in other settings beyond cardiovascular disease prevention where accurate risk calculation is also vital to appropriate clinical decision-making.

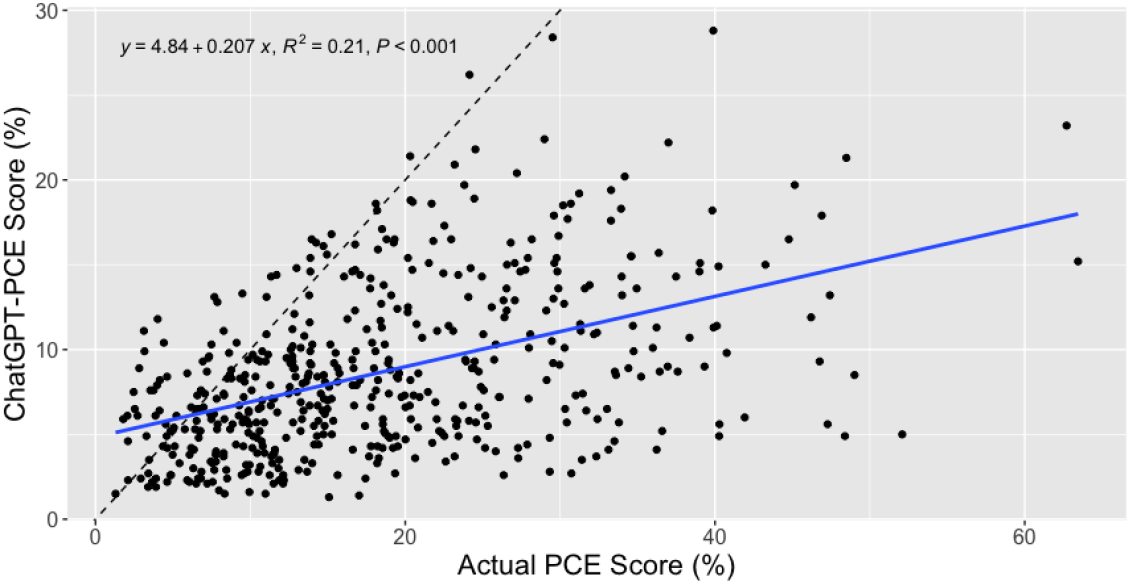

**Figure. Underestimation of true PCE risk estimates (*x*-axis) by ChatGPT (*y*-axis) on synthetic patient data.**

## Introduction

Large language models (LLMs), including general-purpose systems such as ChatGPT as well as more specialized models such as Med-PaLM,^1^ have shown remarkable facility for qualitative tasks in medicine such as question-answering and general clinical reasoning.

Despite the popular conception of ChatGPT and related publicly-available LLMs as mere chatbots, LLMs are in fact capable of tasks beyond question-answering, including arithmetic and mathematical reasoning, albeit with mixed results.^2^ However, the performance characteristics of LLMs for quantitative clinical tasks, including clinical risk prediction, have yet to be assessed. In this preliminary report, we aim to characterize the calibration, reproducibility, and potential sociodemographic bias of ChatGPT-derived Pooled Cohort Equation^3^ (PCE) risk estimates of atherosclerotic cardiovascular disease (ASCVD) as compared to actual PCE risk estimates.

## Methods

Synthetic individual-level data comprising a complete set of PCE predictor variables were randomly generated in R. These synthetic data were used in prompts to generate bulk ChatGPT estimates of PCE risk scores (*ChatGPT-PCE scores*) to assess the 3 domains of performance listed above:

1. *Calibration*: We prompted ChatGPT to generate PCE scores for *n=*500 unique synthetic patients (Prompt C) and compared these ChatGPT-PCE scores to actual PCE scores generated using the R package “PooledCohort”.
2. *Reproducibility*: we generated a new set of *n=*100 synthetic patients, and produced 5 sets of ChatGPT-PCE scores for the same 100 patients by repeating Prompt C 5 times.
3. *Bias*: to assess the potential for bias among sociodemographic lines, we presented ChatGPT with Prompt C to generate ChatGPT-PCE scores for *n=*50 patients, followed by two prompts (Prompts B.1 and B.2) requesting that these 50 ChatGPT-PCE estimates be updated, based on the assumed sociodemographic characteristics of these patients. Overall, this process produced 3 distinct sets of 50 ChatGPT-PCE scores, with each set corresponding to an assumed sociodemographic context.

ChatGPT-3.5 (9 May 2023 version) was used for all experiments. Calibration was assessed graphically and via the Pearson correlation coefficient, while analysis of variance (ANOVA) was applied across repeated rounds of prompting to test for changes in scores across rounds in the reproducibility and bias experiments. The text of Prompts C and B.1/B.2, together with representative R code, are provided in the Appendix.

## Results

Compared to the actual PCE estimates for the 500 synthetic patients (**Figure 1**), calibration of the ChatGPT-PCE scores appeared poor. The ChatGPT-PCE score consistently under-predicted actual PCE scores, although the two sets of scores exhibited modest correlation (Pearson correlation coefficient 0.46, *p* < 0.001). Moreover, individual ChatGPT-PCE scores, when re-generated 5 times with identical synthetic data, did not appear reproducible, being significantly different across 5 attempts for each individual synthetic patient (one-way ANOVA *p* = 0.010) (**Figure 2**). Finally, when prompted to update a set of risk estimates based on whether the data were assumed to derive from a safety-net clinic (Prompt B.1) or from a clinic in an affluent suburb (Prompt B.2), ChatGPT-PCE risk estimates were revised significantly upwards, then downwards, respectively (one-way ANOVA *p* < 0.001) (**Figure 3**).

**Figure 1.**
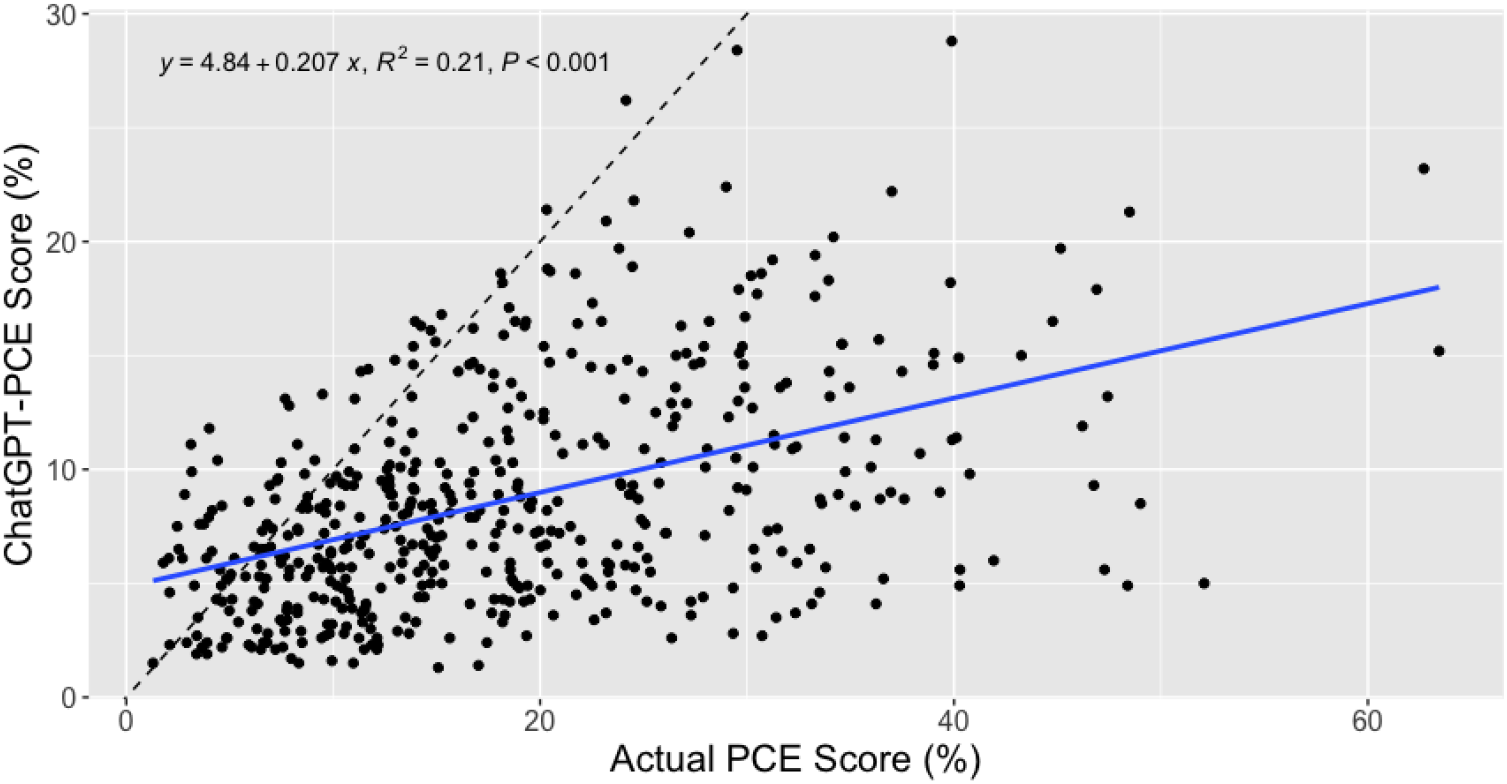
Comparison of Pooled Cohort Equation (PCE) scores generated by ChatGPT (ChatGPT-PCE scores) to true PCE scores on individual synthetic patients. The blue line depicts the best-fit line, and the statistics in the upper left corner are those associated with that line. The dashed line depicts the 45-degree line associated with perfect calibration.

**Figure 2.**
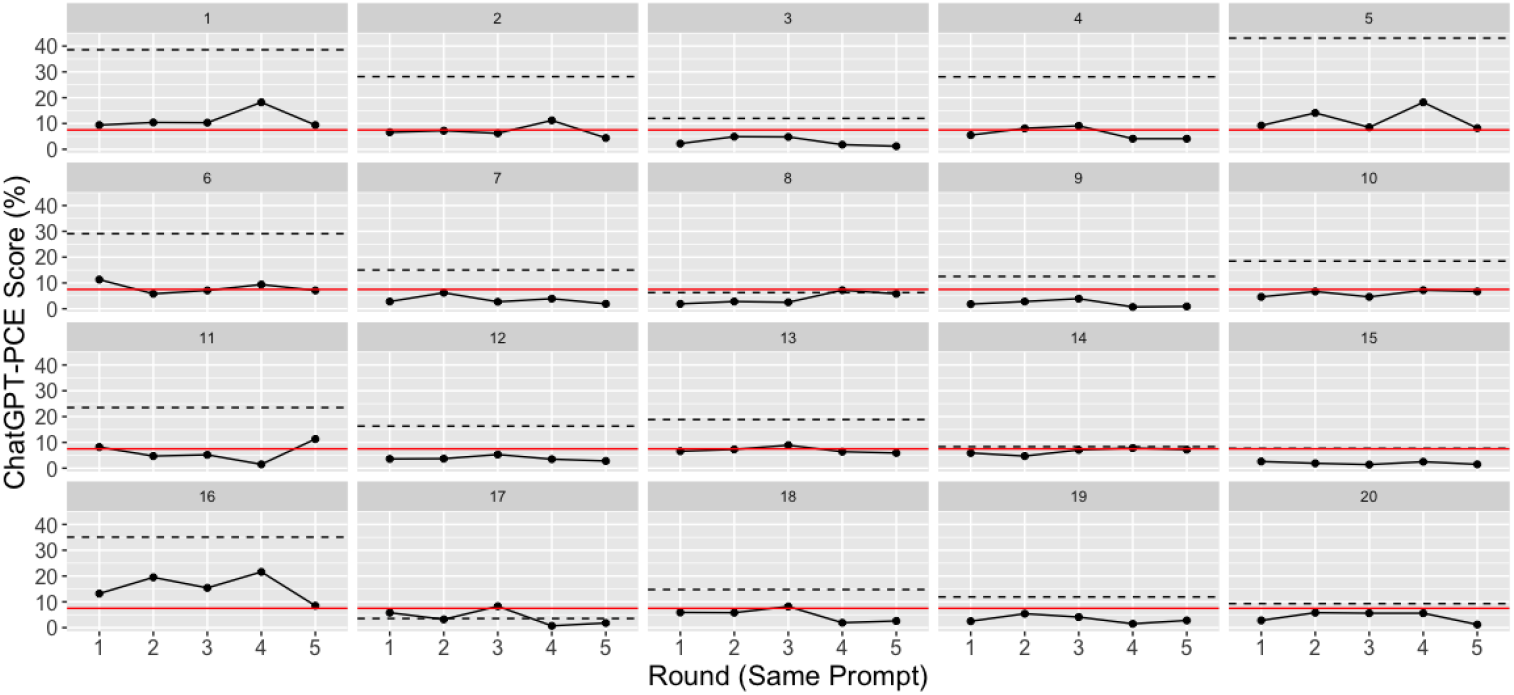
Reproducibility of ChatGPT-PCE estimates for a subset (20 shown) of the n=50 synthetic patients. Each panel corresponds to one synthetic patient, and with points denoting their ChatGPT-PCE scores generated across 5 rounds of prompting. The dashed line denotes the true PCE risk estimate for that patient, while the red line denotes the 7.5% PCE threshold. Both substantial variability in ChatGPT scores and frequent reclassification with respect to the 7.5% threshold are observed.

**Figure 3.**
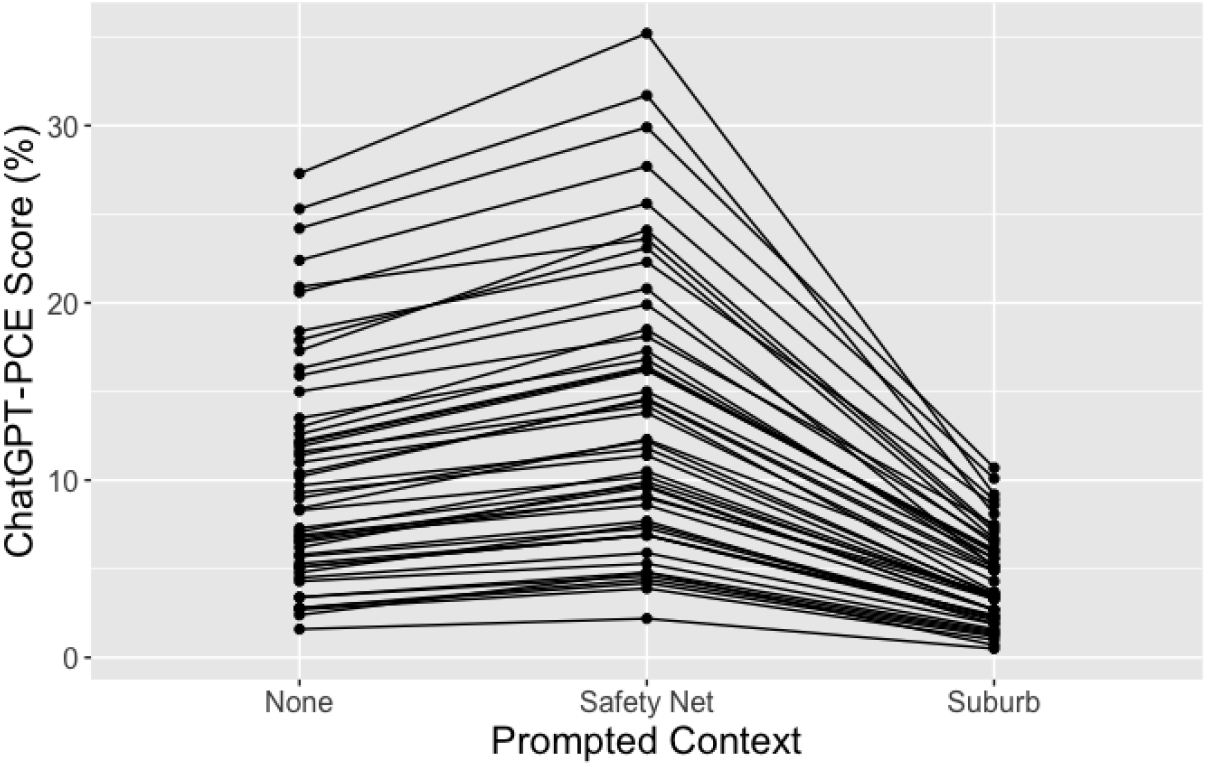
Sensitivity of ChatGPT-PCE scores to additional context potentially indicative of sociodemographic status of synthetic patients (*x*-axis). Each point depicts the ChatGPT-PCE risk estimate for that patient under three different sets of context. The data were initially generated with no such context (“None”), then were updated under the assumption that the patients were treated in a safety-net clinic (“Safety Net”; Prompt B.1) then again updated under the assumption they derived from a clinic located in an affluent suburb of a Midwestern city (“Suburb”; Prompt B.2).

## Discussion

This study found that ChatGPT produced poorly calibrated, and individually highly variable, estimates of ASCVD risk compared to those obtained via the true Pooled Cohort Equations. However, despite the demonstrated propensity of LLMs to “hallucinate” (fabricate) output, the ChatGPT-PCE estimates did exhibit significant correlation with the true PCE scores. Moreover, ChatGPT-PCE scores, upon re-prompting, appeared sensitive, and arguably unnecessarily so, to contextual indicators of patient sociodemographic status. Higher ChatGPT-PCE scores were generated for patients assumed to be treated at a safety-net clinic, while the same set of patients, this time assumed to be treated in a clinic in an affluent suburb, received far lower scores. These adjustments appeared to be performed in an idiosyncratic manner with no apparent justification (e.g. an adjustment factor, equation, or re-calibrated model) behind why individual scores were adjusted as observed.

Our study design presented ChatGPT with synthetic patient data examples to generate the ChatGPT-PCE risk estimates. Here, our approach relying on synthetic data forecloses the possibility, however improbable, that ChatGPT had simply memorized these particular data.

Nevertheless, it is not immediately clear why the ChatGPT-PCE scores appeared to carry at least *some* information about true PCE risk scores, given the moderate level of correlation observed between these two sets of scores. Further work remains to probe ChatGPT and other LLMs to understand the origins of this observation.

Our study, while preliminary, has several limitations, which also present avenues for further work. First, it remains to be seen whether our results can be replicated by other LLMs, including ChatGPT-4, Anthropic’s Claude 2, and Google’s Bard, among others. Second, our prompt may not necessarily reflect how a LLM would be used to generate risk estimates in practice. Indeed, it may not be immediately clear why a LLM would be needed to generate risk estimates at all, given that risk calculators already exist and are readily available.

However, insofar as PCE risk estimates remain integral to decision-making for primary prevention of ASCVD,^4^ their accurate calculation is essential for systems interfacing with, and reasoning based on, patient data from encounters for ASCVD prevention. Many other settings beyond ASCVD prevention also depend on accurate risk estimation for appropriate clinical decision-making, as well. Future work could assess LLM performance based on patient vignettes or prompts more reflective of actual practice. Ultimately, LLMs may rely on the ability to hook into an external source, such as a “code interpreter”,^5^ to interface with the appropriate risk calculator and directly compute the desired estimates. However, no such interfaces for clinical risk calculators yet exist, and so such abilities remain untested.

Given current efforts towards LLM-electronic health record integration, our preliminary findings may have broad implications. In particular, our finding that ChatGPT-PCE estimates carried at least some information regarding true estimated risks is surprising. From a safety perspective, this finding may also be concerning insofar as it demonstrates the potential for automation bias^6^ engendered by inappropriately-calibrated trust in quantitative output that ostensibly appears correct.^7^ Altogether, further work remains—not only to build on our preliminary results, but also to characterize the performance of LLMs on a wider variety of clinical risk calculators and to investigate methods with potential to improve their performance on these and related tasks, including chain-of-thought prompting^8^ and other approaches to prompting.

## Data Availability

The data produced in the present study are available upon request.

## Appendix to

### Example Prompts

#### Prompt C

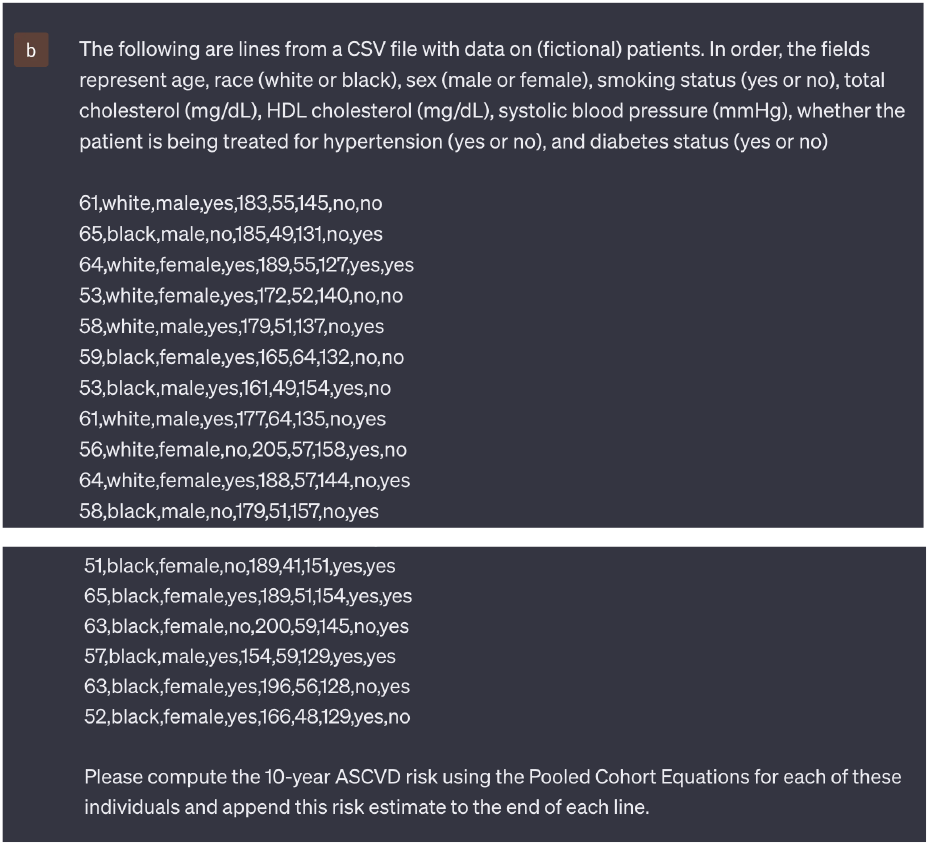

#### Prompt B.1

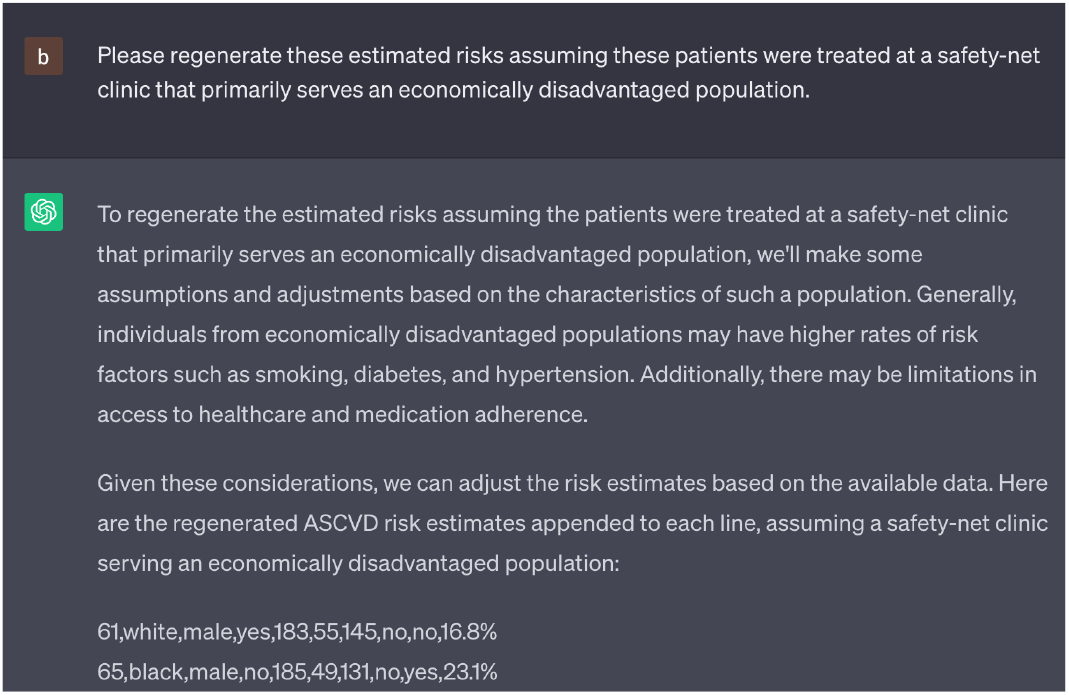

#### Prompt B.2

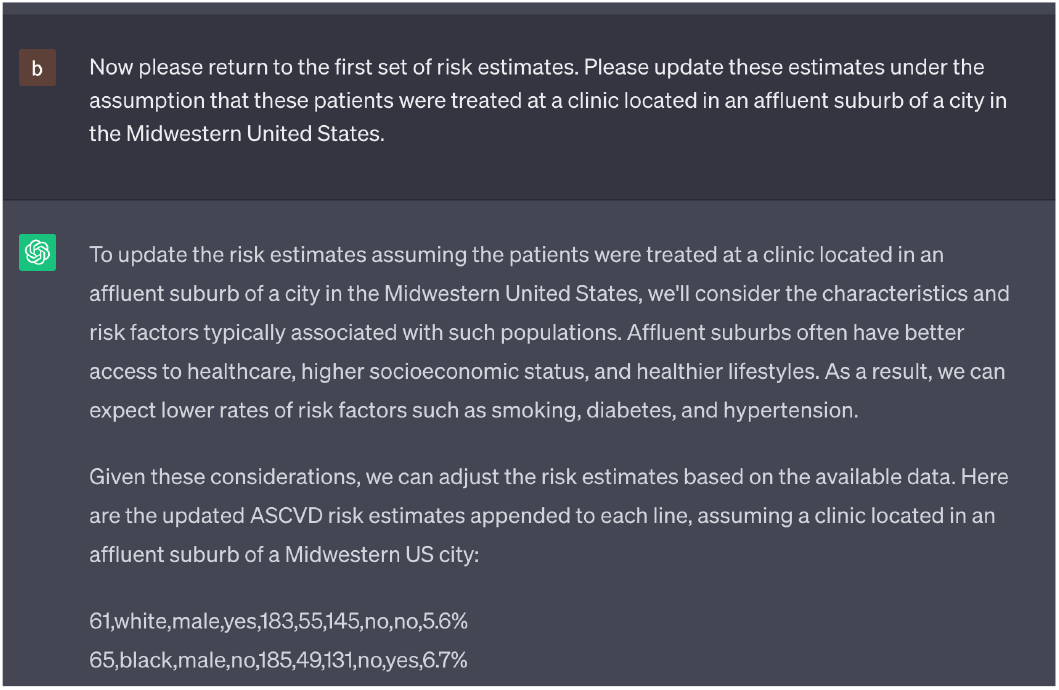

#### Example R code

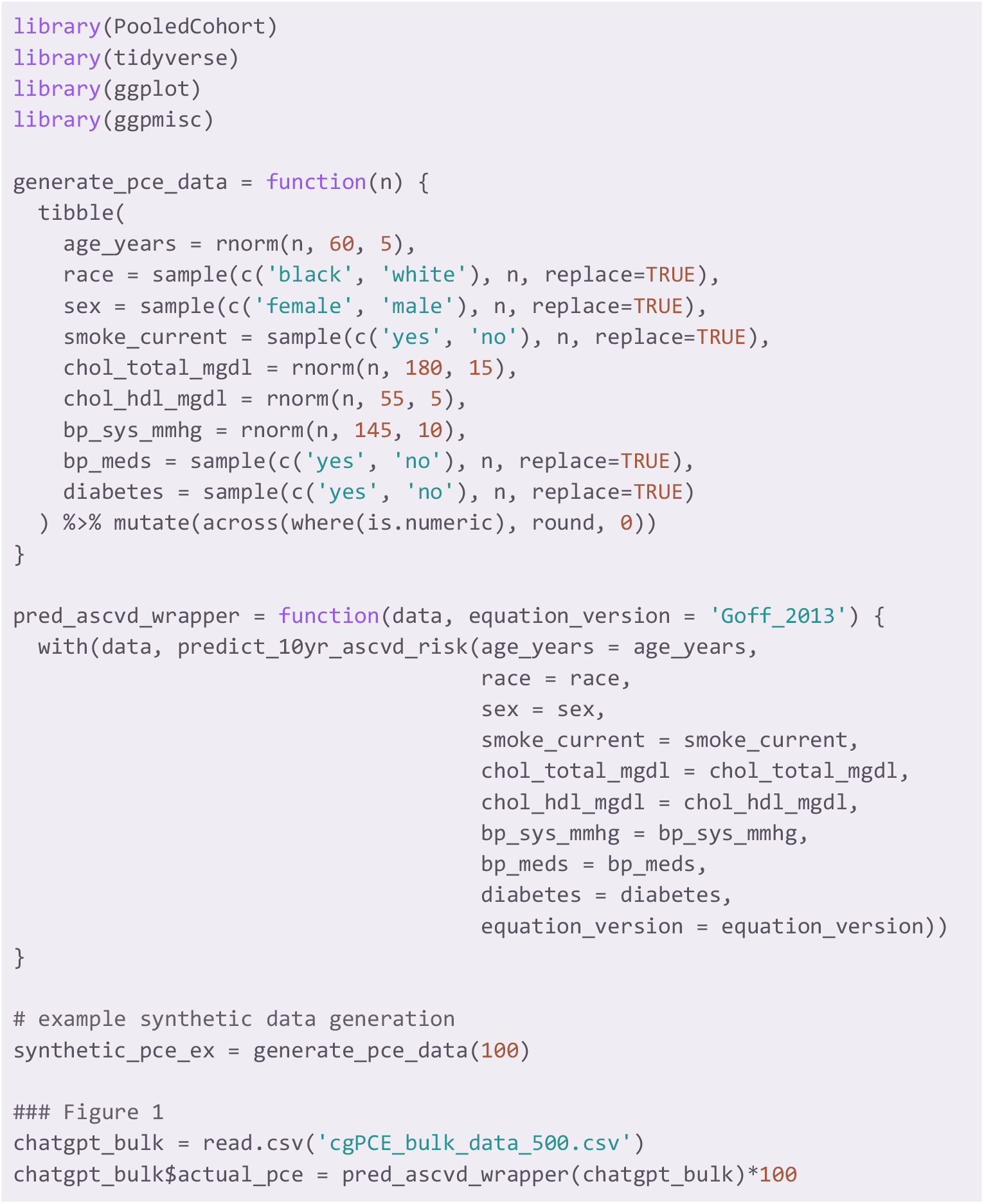
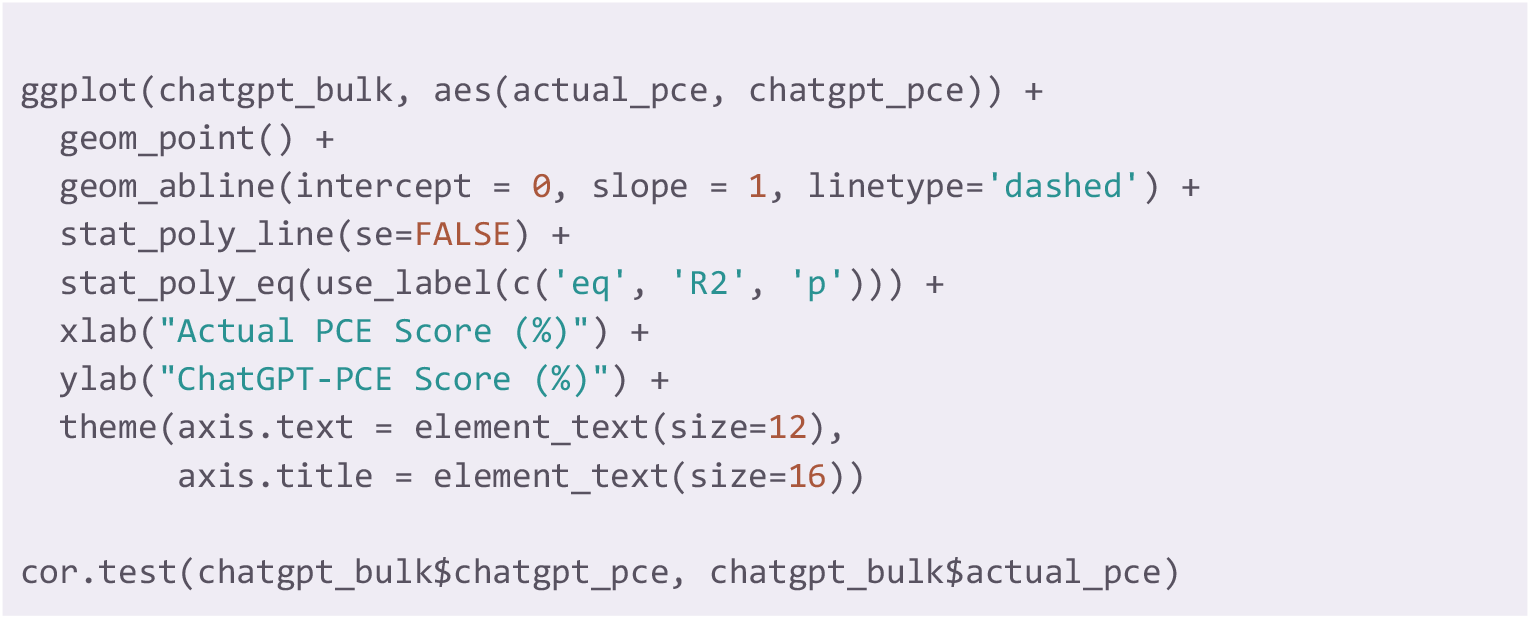

